# Health Exposure Records and Occupations (HERO) Summary: Development of Occupational Exposure Summary for clinical utility in Military Populations

**DOI:** 10.1101/2024.10.03.24314842

**Authors:** Immanuel B.H. Samuel, Kamila Pollin, Sherri Tschida, Lily Reck, Alan Powell, Jessica Mefford, Jamie Lee, Teresa Dupriest, Michelle Prisco, Stephen Fischer, Jose Ortiz, Robert Forsten, Charles Faselis, John Barrett, Matthew Reinhard, Michelle Costanzo

**Affiliations:** The War Related Illness and Injury Study Center, Washington, DC, USA; Complex Exposure Threats Center of Excellence, Department of Veterans Affairs, Washington, DC, USA; Big-data Research Artificial Intelligence and Neuroscience Lab, Washington, DC, USA; The Henry M. Jackson Foundation for the Advancement of Military Medicine Inc, Bethesda, MD, USA; Department of Psychology, Georgetown University Medical School, Washington, DC, USA; Department of Medicine, Uniformed Services University, Bethesda, MD, USA; Washington VA Medical Center, Washington, DC, USA; Explosive Ordnance Disposal Information Management System (EODIMS), Air Force Civil Engineer Center, Functional Management Office (FMO) (AFCEC/CBFP), Joint Base San Antonio, TX, USA; Department of Psychiatry, Georgetown University Medical School, Washington, DC, USA

**Keywords:** Exposure, Summary, Clinical Needs, Military Exposures, Veterans

## Abstract

**Introduction:** Military exposure summarization is critical for Veterans with complex environmental, occupational, or toxic exposures. Existing methods are limited by technical language, incompatible data formats, and difficulty in prioritizing information. Clinicians require concise, standardized, and easily interpretable exposure summaries to facilitate rapid assessment. This report is part of a broader programmatic effort to collate military exposure information from established, as well as new sources of data to improve VA exposure-informed healthcare.

**Methods:** In a collaborative effort, VA clinicians from multiple specialties participated in a structured clinical needs assessment interview to identify the most clinically useful information to be included in the Health Exposure Records and Occupations (HERO) summary. The interviews covered summary length, exposure prioritization, demographics, military occupational history, features characterizing exposure, resilience factors, health outcomes, and impact on clinical practice.

**Results:** Consensus recommendations prescribed a concise summary with clear language, basic military demographics, and critical military exposures that prioritize exposures that require further investigation. Based on recommendations, the HERO summary also includes types of exposure, proximity, route, symptoms at the time of exposure, exposure period, duration, frequency, and protective controls used.

**Conclusion:** This perspective piece not only assesses the clinical need for exposure summarization and the optimal format for the HERO summary, but also highlights its potential impact. The HERO summary, as a tool, offers improved time efficiency, consistency in exposure-informed care across the VA, enhances communication between Veterans and providers, and improves understanding of the association between military exposures and health outcomes, potentially transforming the VA healthcare system.

## Introduction

Veterans can require specialized clinical services to meet their post-deployment health needs and recent focus has been oriented towards the development of exposure-informed care across the VA enterprise. This requires clinicians to not only understand military service and exposure information but also apply that to clinical recommendations and overall care. The challenge is that although military environmental exposures (MEE) and service records offers a critical source for improved understanding and management of complex occupational health concerns, this information often lacks continuity with Veteran health data. A comprehensive method for understanding exposures would thus not only include the capacity to structure MEE information to address research questions but would also offer concise clinical summaries that could aide VA providers in delivery of exposure-informed care. To achieve this, the first step requires the consolidation of complex military environmental exposure records into exposure common data elements (ExCDE) based on occupational histories as proposed in the Linked Exposures Across Databases (LEAD) framework (Samuel et al., 2024). Next this aggregate knowledge must be summarized in a way that clinical utility- for example being easily understood across varying levels of exposure literacy, succinct for exposure review in clinics and aligned with health outcomes. This perspective piece describes the clinical needs assessment that were attained through interviews of a group of military healthcare leaders, which resulted in the development of the Health Exposure Records and Occupations (HERO) Executive Summary. HERO offers the clinical capstone that connects exposure extraction using LEAD (Samuel et al., 2024) to clinical implementation thereby establishing a scalable answer to the implement of VA exposure-informed healthcare.

HERO was created based on the judgement of the authors that a clinical summary significantly augments the impact of LEAD tiered data aggregation method that is intended to consolidate MEE information (Samuel et al., 2024). Prior application of the LEAD framework involved demonstration of data aggregation techniques from military incident records that have previously never been utilized for health applications. Specifically, LEAD was able to connect data from military incident reports stored in Explosive Ordnance Disposal Information Management System (EODIMS), with information in the Individual Longitudinal Exposure Record (ILER) that contains more traditional methods of objective exposure assessments, microenvironmental samples and personal monitors (Lioy et al., 1995; Wambaugh et al., 2019) as well as with subjective histories from Veteran Military Occupational and Environmental Exposure Assessment Tool (VMOAT) (Samuel et al., 2024).

While LEAD enables extraction of ExCDEs (Level 2) from raw data (Level 1) to support research oriented statistical analysis, such information also needs to be made available as a concise exposure health summary that could help clinicians understand a Veteran’s exposure profile (Level 3). This report describes the structured interviews that were completed to generate a display template of Health Exposure Record and Occupations executive clinical exposure summary (Level 3) using mock data that is reflective of a Veteran within the Explosive Ordnance Disposal (EOD) career field to illustrate a complex military career history.

## Methods

Clinicians with expertise in military medicine were interviewed about their requirements for an exposure report and how it can help improve care for Veterans. Web-based interviews were transcribed with ten subject matter experts, including five physicians (specializing in preventive medicine, occupational and environmental medicine, epidemiology, family medicine, and psychiatry), two psychologists, two health coaches, and one nurse with an extensive exposure assessment background. Eight of our subject matter experts are Veterans, which adds military environmental exposure knowledge to the answers provided. The structured interview was sectioned into eight parts. Questions included:

1. Assessing the need for an exposure summary, for example: If you had a history of all the places that a Veteran has been and the exposures that they potentially had given their job duties, concisely summarized, would that be beneficial?
2. Optimal exposure summary length: What would be ideal for a concise Veteran military exposure summary report?
3. Demographics information, for example: Is there a need to include information on the Veteran’s socioeconomic, environmental, or pre-service factors that could influence their experiences during military service?
4. Military history, for example: Given the availability of the DD214 form, should this exposure report include military service history?
5. Prioritization of Exposures, for example: Should we address complex exposures? How do you define a complex exposure?
6. Features characterizing exposure, for example: Would you like to see the kind of features of that exposure, like proximity route symptoms, duration, frequency, age, and other hierarchy of controls?
7. Resilience and health outcomes, for example: Should the report consider potential resilience factors for exposures like supportive relationships and coping mechanisms that could mitigate the effects of exposures on outcomes?
8. Impact of exposure summary on clinical practice, for example: How would the exposure report help Veterans as they engage with their clinicians and review their own exposure history and treatment progress?

The following is a summary of the feedback obtained during the interviews.

## Results

### Part 1: Assessing the Need for an Exposure Summary

All respondents expressed a need for a concise exposure summary, emphasizing how such a summary would improve their clinical processes. Such a need stemmed from the inadequacy of current methods of military exposures assessment, which are varied, complex to access, and laborious to capture. Useable exposure data should provide consistent interpretation of extensive exposure information. Issues on information access rights and the education necessary to gain rights and use systems (for example, the Individual Longitudinal Exposure Record, ILER) were additional barriers expressed by the respondents. Furthermore, extensive exposure education is needed to work with information systems and although the War Related Illness and Injury Study Center (WRIISC) provides educational briefings with regularity, the specifics of each case and manipulation of differing source locations further increases the time required to perform a clinical review. To address these issues, respondents agreed that an easily accessible exposure summary that provides a concise, high-level overview of multiple exposures could help streamline exposure assessment processes for VA providers by prioritizing relevant exposure information that could result in improving patient care, enhancing communication between healthcare professionals, and ensuring better understanding and addressing of concerns related to military exposures.

One example: A respondent provided a real-world scenario where the patient was in pain on admission to the clinic. The clinician said since this scenario does not give ample time for a detailed exposure review, it is not appropriate to ask the patient to provide their exposure history when they are in pain and facing acute health complications. In this scenario, a concise 1-page summary would be incredibly beneficial in understanding the patient’s history. Such a document given to clinicians can provide a greater understanding of the Veteran and help focus resources on caring for the acute health condition.

### Part 2: Report Length and Format

Most respondents agree that an ideal exposure summary should be one to two pages long, with some suggesting a maximum of one page for brevity and ease of reading. When interviewing the Veteran in the clinic, the summary should prioritize key exposures and concerns for further investigation by the provider. It should use structured and standardized terminology, only abbreviations if explained in detail, and should be written in language that is easy to understand. Some suggest using bullet points or graphs to present information concisely without exceeding one page. The summary should provide a clear and digestible overview of the critical military exposures, allowing clinicians to grasp the main points quickly within one to two minutes.

### Part 3: Demographics Information

All except one respondent agreed that basic demographics such as age, sex, and race are essential for understanding the Veteran’s background and experiences during military service. Basic demographic information is necessary to anchor the interpretation of the exposure information. While one respondent expressed that such information can be accessed from medical records, other respondents suggested that having basic demographic information within the report helps cut down the additional time required to pull up the patient’s medical record to access that information. Clinicians with psychology, psychiatry, and health coaching backgrounds also expressed interest in socio-economic, environmental, childhood, and other pre-service factors. One suggested including Department of Defense (DoD) ID, Birthdate, and social security number (SSN). Others suggested that only basic demographics are necessary for the executive summary, and additional social factors and cultural differences may be extracted from electronic health records or captured in a separate, specialized report. There was also concern that including pre-service factors in the executive summary might unintentionally affect the Veteran’s disability rating, so it is prudent that the inclusion of such information in any supplementary material must highlight the intended use case as accompaniment to medical histories, rather than disability and service connection assessment.

### Part 4: Military History

Most respondents expressed the need to include enlistment year, total years of service, period, and service location. The interviewees noted the importance of the nature of their occupational assignments, including unique exposure(s) associated with duties, branch of service, highest military rank achieved, and significant participation in conflicts and operations. These deployments include combat deployments overseas and non-combat deployments inside the Continental U.S. (CONUS) or outside the Continental U.S. (OCONUS). Chronological military history was stressed as a crucial element in assessing the cumulative nature of exposures. Some respondents also expressed interest in including military training history, pre-service traumatic experiences, such as contact sports history, and other traumatic stress history, stating that can be “a big factor in their [Veteran’s] ability to move forward with healthy lifestyle choices.” One respondent also suggested including information on special pay or medals awarded. These reports should accompany disclaimers since they may be incomplete or not reflect actual service history - for example, for some national disasters, records may be incomplete if they are not associated with special pay. Others said that they do not find branches of service, specific ships/bases/units, and other training experiences particularly useful in their clinical practice.

### Part 5: Exposure Prioritization

Exposures should be prioritized based on their potential health impacts and complexity. High-risk exposures with clear links to specific processes or syndromes should be prioritized, followed by complex exposures with multiple contributing factors. The following are some of the exposures respondents brought up to be considered for prioritization: Chemical exposure incidents (e.g., Agent Orange), radiation events, burn pits, environmental hazards, infectious diseases, blast and traumatic stress events, electronic countermeasures, occupational exposures associated with certain health conditions, military sexual trauma, prolonged noise exposure, high alert psychological stress, physical stress, potential effects of medications or vaccines taken during deployment, and depleted uranium exposure. Prioritization should also take into account the duration and intensity of exposures, as well as the potential for interaction between simultaneous exposures. Respondents suggested that a one-page report highlighting multiple exposures can be a starting point for further discussion with patients and healthcare providers. Two respondents indicated that since complex exposures are highly varied and constantly evolving with military technology, a predefined list of complex exposures could be developed through expert review and literature review to guide the prioritization process in exposure reports. Additionally, one respondent noted that providing all the exposures recorded across various databases in the summary increases the report’s length and might result in unwarranted exposure-related concerns. It may be beneficial to prioritize specific exposures based on known health outcomes.

### Part 6: Exposure Characterization

Since exposure characterization is not standardized, the ExCDEs developed by the LEAD framework were used in this part of the interview. All respondents agreed that exposure features such as proximity, route, symptoms at the time of exposure, exposure period, duration, frequency, and type of protective equipment worn were important in characterizing exposures. Additionally, respondents suggested other features relevant to exposures, such as the potential for chronic effects, exposure to physical state (powder, liquid, gas), and medical treatment sought or received following exposures. Most tertiary care clinicians favored including a detailed characterization of exposures. However, one primary care clinician suggested that while these features may be relevant to exposure characterization, such details may be too verbose for inclusion in a concise executive summary and suggested inclusion in the supplementary. This thought was reflected by other clinicians who agreed that detailed information may not be relevant for such clinicians as frontline providers who may need more exposure education to utilize such information. Respondents agreed that a composite exposure score in the executive summary with adequate documentation in the supplement material on how the score was obtained would be helpful, especially for those who need more exposure education to interpret the nuances.

### Part 7: Resilience factors and health outcomes

Respondents suggested including supportive relationships and coping mechanisms to highlight the individual’s ability to mitigate the effects of exposures on their mental and physical well-being. These factors can provide a comprehensive view of the Veteran’s overall health and better inform treatment approaches, especially for health coaches and psychologists. However, such data may not be readily available and thus could be sparse. There was more agreement in including common chronic conditions such as diabetes and hypertension, expected military service-related outcomes such as traumatic brain injury and post-traumatic stress disorder, and lifestyle factors like smoking status, alcohol use, and body mass index. While one respondent recommended a complete medical history, another suggested only having a predefined list of conditions to include to maintain brevity. It is recommended to briefly mention these health outcomes in the executive summary if categorized succinctly, and ICD codes are avoided. A few respondents expressed that the summary should primarily focus on exposures without including extensive details about resilience factors and health outcomes to keep the report concise and focused on the primary purpose of assessing exposure risks. Other respondents warned that listing all health outcomes on the exposure report may lead Veterans to draw premature conclusions, causing unnecessary concern.

### Part 8: Impact of Exposure Summary on Clinical Practice

The respondents shared their opinions on the usefulness and potential benefits of having a concise exposure summary for Veteran care. Most respondents agreed that such a summary would be beneficial, as it could save time, improve consistency across clinics, facilitate better communication between patients and healthcare providers, and aid in understanding the association between exposures and potential health outcomes or treatment progress. A respondent stated, “One of the most important things is to not have a lot of variability” and “we need to measure [exposures] with consistency and have a consistent framework.” The summary should be a reference tool for discussing past exposures and current health issues. Respondents shared that it should be user-friendly for Veterans and their medical providers and that accessibility of the same report across VISNs is critical for clinical utility. However, some also acknowledged that a summary might be misinterpreted if Veterans do not understand it correctly without prior knowledge of exposures.

## HERO Summary Guidelines and Template

The responses gathered from the interviews provided various viewpoints on clinical needs for a concise exposure summary, its contents, impact, and areas to exercise caution. While some views differed across respondents, the following consistent recommendations should be considered when developing exposure summaries meant for clinical use:

1. Be easily accessible for all providers
2. Not exceed 1 page
3. Contain a chronological list of military service with details such as location, duration, and duties
4. Prioritize exposures based on established information relating exposures to health outcomes
5. Characterize exposures based on proximity, route, symptoms immediately following exposures, period of exposure, duration, frequency, and usage of preventive controls, provided these factors can be represented in a concise manner
6. Capture chronic health outcomes, outcomes typically related to military service, and lifestyle factors commonly assessed during VA clinical intake, only listing medications and therapies if they can be concisely expressed
7. Provide composite scores with adequate documentation of how it was calculated
8. Reduce technical jargon, abbreviations, and the use of coding systems like ICD to make it easily readable for both clinicians and Veterans
9. Include disclaimers and caveats to highlight that information may be incomplete and may not sufficiently reflect Veterans’ actual military experience and that Veteran self-reports must accompany such summaries
10. Ensure that the summary’s intended use case is to provide a starting point for further detailed clinical review and that it is not meant to be used for disability assessment or service connection determination

Based on the interview responses, the need for an exposure summary is evident based on the interviews provided by clinicians with different professional backgrounds. An executive summary would improve exposure review and care, particularly in frontline healthcare settings. In addition, accessing an overview of a patient’s exposure history enables clinicians to better connect to the patient, thus enhancing emotional care and concordance between provider and Veteran (Bloeser et al., 2024). Based on the above set of guiding principles and in collaboration with the EOD community, we provide a template of the HERO summary based on mock data that reflects the typical military occupational history, exposures, and health outcomes of an EOD Veteran (Figure 2; PDF included as part of the supplementary material).

**Figure 1:**
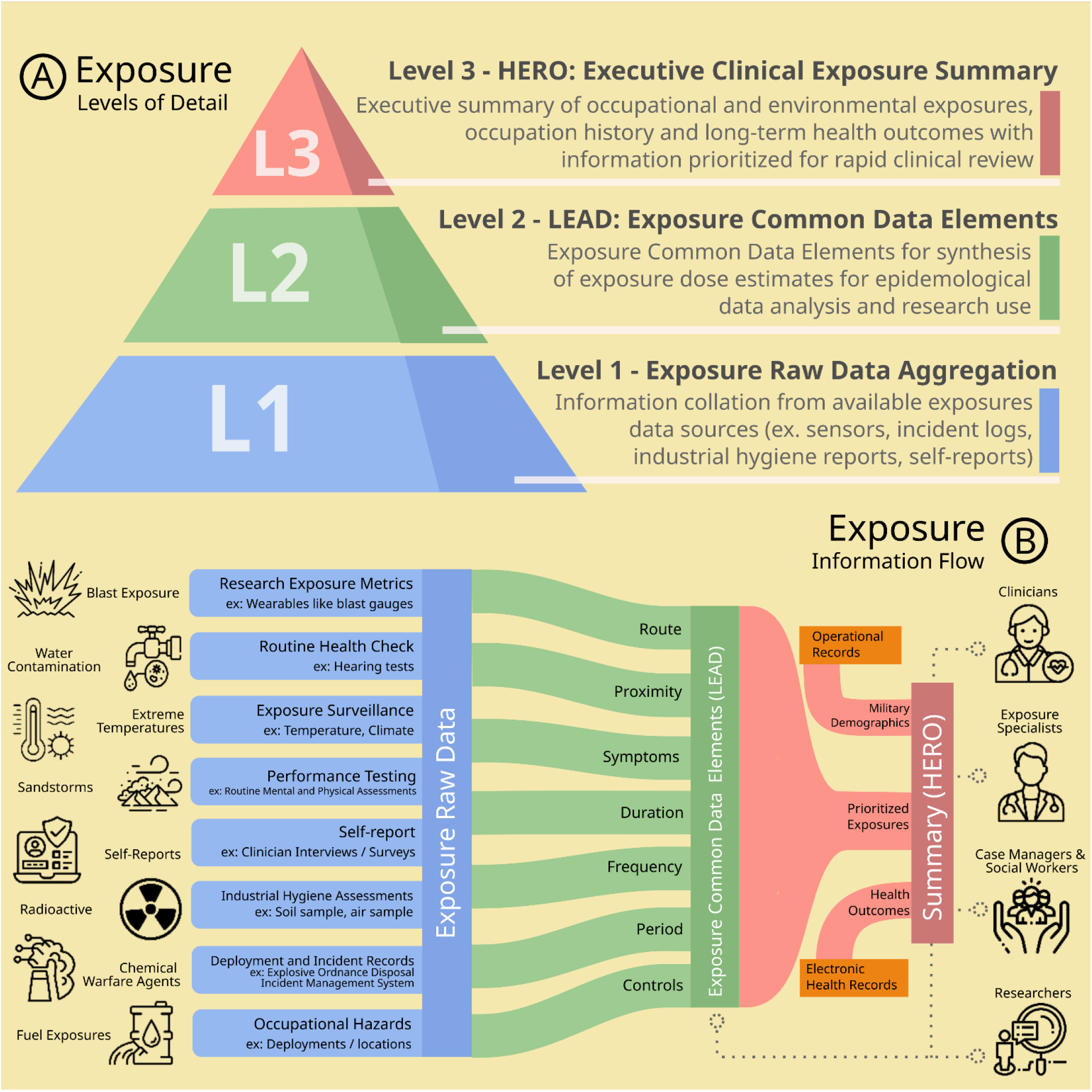
Exposure data levels of detail and information flow: Sparse varying information from multiple data sources need to be aggregated in a detailed manner (Level 1). The raw exposure data needs to be further homogenized using the Linked Exposures Across Databases (LEAD) framework (Level 2). For clinical use, the Health Exposure Records and Outcomes (HERO) summary aims to provide a concise exposure summary to facilitate rapid clinical exposure review (Level 3)

**Figure 2:**
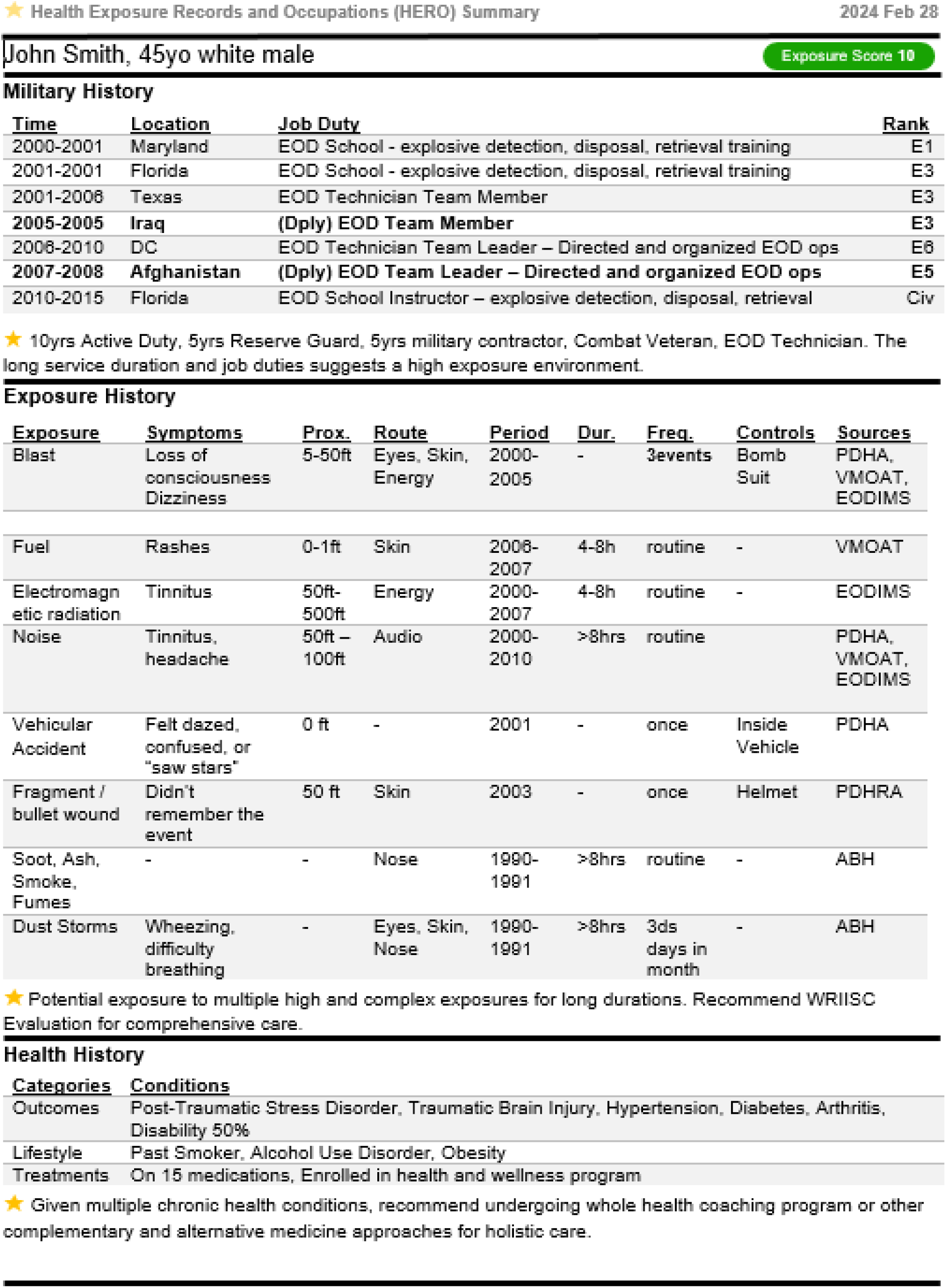
Health Exposure-Records and Occupations (HERO) Summary template.

The template from the mock data demonstrates how a concise, easy-to-read summary of military exposures can aid in efficient clinical decision-making, streamline communication between patients and healthcare providers, and facilitate a better understanding of the potential impacts of exposures on Veterans’ health. Although considerable research has focused on improving understanding of environmental health risks, including development of computational exposure tools and predictive model risk assessments, such methods are highly variable, and the exposure results often are not consolidated in succinct, easy-to-understand summaries to facilitate clinical care. Variations in the exposure information and assessment methodologies result in varied exposure reports that are hard to interpret without an approach that integrates this knowledge into meaningful clinical insights. Furthermore, existing methods are limited by technical language, incompatible data formats, and difficulty prioritizing information.

The critical recommendations gathered during the interviews emphasize the importance of prioritizing exposures based on established associations with health outcomes, concisely characterizing exposures, including relevant chronic conditions and lifestyle factors, providing composite scores with adequate documentation, reducing technical jargon, abbreviations, and ensuring that the summary is accompanied by disclaimers and caveats highlighting its limitations.

Such exposure summaries could significantly improve clinical care for Veterans by providing healthcare providers with essential information about a patient’s military service and exposures in a concise format. The HERO summary can serve as a starting point for further detailed clinical review, allowing clinicians to focus on caring for their patients’ acute health conditions while also considering any potential long-term effects of past exposures. By making this information accessible, healthcare providers could provide more personalized and comprehensive care for Veterans, ultimately improving their overall health outcomes.

## Future directions

While this report only includes a template of the HERO Executive Summary, we intend to generate case reports based on participant data from ongoing research studies at WRIISC. Additionally, we aim to use the interview responses to develop a short, focused web survey to expand the participant pool and get feedback from a broader range of clinical backgrounds, such as primary care, specialty care and junior vs. senior clinicians. The feedback from the responses will be incorporated into the HERO template. Furthermore, we aim to share the tools developed in this report in education and training material for clinical care providers as part of the WRIISC education webinar series to improve exposure literacy and facilitate consistency in exposure information capture and interpretation.

With the VA’s commitment to addressing military environmental exposures in Veterans, VA providers need a comprehensive summary in which the HERO Executive summary can provide. This tool can be integrated into Veteran’s electronic health records or other VA exposure platforms helping tailor medical care to each Veteran’s specific risks related to military environmental exposures.

## Conclusion

Existing summarization methods face challenges in determining which information is relevant or essential for VA expose-informed care. Automated electronic health record (EHR) summarization faces challenges in detecting redundancy, describing temporality, determining salience, accounting for missing data, and utilizing encoded clinical knowledge [Pivovarov2015]. Currently, exposure data aggregation in the VA and DoD is done by the ILER, which are intended to be electronic records of exposures. While such a unification of exposure information is critical to improving exposure assessment, ILER reports are extensive and can be difficult to understand. This can pose a challenge, particularly in clinical contexts where rapid evaluation and interpretation are essential. While exposure literacy can be high in tertiary care settings (such as in WRIISC and Complex Exposure Threats Center of Excellence - CETCE), it can be lacking in frontline care settings where time is critical factor when delivering services to Veterans. Furthermore, exposure assessments from various databases like Post Deployment Health Assessments (PDHA), Periodic Occupational and Environmental Monitoring Summary (POEMS), Joint Longitudinal Viewer (JLV), and Defense Occupational and Environmental Health Readiness System- Industrial Hygiene (DOEHRS-IH) contain data in different formats due to variations in survey length, complexity as well as different intended use for such data. HERO framework provides a method to aggregate, condense, and prioritize information improving readability and interpretability-key factors in routine clinical practice.

## Data Availability

Only mock data is used in this perspective report

## Acknowledgments

None to report

## Contributions

IS: Original draft, conducted Interviews and analysis KP, LR: Review and analysis.

AP, JM, JL, TD, ST, MP, SF, JO, RF, CF, JB, MR, MC: Final review

